# Clinical Usefulness of Artificial Intelligence–Enabled Electrocardiography for Pulmonary Hypertension Detection: A Meta-analysis Based on Pre- and Post-Test Probability

**DOI:** 10.64898/2026.10.25.26365896

**Authors:** Johann Edjimbi, Ayse Elif Kesaf, Alyssa A. Grimshaw, Aryan Gajjar, Krishna Allam, Hooman Bakhshi

## Abstract

Pulmonary hypertension is frequently diagnosed late because its early manifestations are nonspecific. Artificial intelligence-enabled electrocardiography may provide a scalable method for identifying patients who warrant further evaluation. We searched eight databases from inception through May 2026 without language restrictions for studies evaluating artificial intelligence or machine-learning algorithms applied to standard 12-lead electrocardiograms against invasive hemodynamic or echocardiographic reference standards. Two reviewers independently selected studies, extracted data, and assessed risk of bias. Sensitivity and specificity were pooled using a bivariate random-effects model. The protocol was prospectively registered in PROSPERO (CRD420251149532). Five retrospective studies comprising 103,884 individuals were included. Pooled sensitivity was 0.81 (95% CI, 0.79-0.82), pooled specificity was 0.80 (95% CI, 0.76-0.83), and the summary area under the curve was 0.85 (95% CI, 0.81-0.88). The positive likelihood ratio was 4.03 (95% CI, 3.38-4.80), and the negative likelihood ratio was 0.24 (95% CI, 0.23-0.26). At a pretest probability of 20%, positive and negative results corresponded to posttest probabilities of approximately 50% and 6%, respectively. Heterogeneity was substantial, but leave-one-out analyses did not materially alter pooled estimates. Artificial intelligence-enabled electrocardiography showed good performance for detecting pulmonary hypertension and may support targeted case finding. Prospective studies should determine whether its use improves diagnostic pathways and patient outcomes.

**Author summary:** Pulmonary hypertension is a serious condition that is often recognized only after symptoms have progressed. The standard electrocardiogram is inexpensive and widely available, but visual interpretation alone may miss early or subtle changes. We examined whether artificial intelligence can improve the ability of the electrocardiogram to identify pulmonary hypertension. We combined results from five studies involving more than 103,000 people. The artificial intelligence models correctly identified approximately 81% of people with pulmonary hypertension and correctly classified approximately 80% of people without it. In a setting where one in five patients has pulmonary hypertension, a positive result would raise the estimated probability to about one in two, while a negative result would lower it to about one in sixteen. These findings suggest that artificial intelligence-enabled electrocardiography may help select patients for echocardiography or invasive testing. It should not be used alone to establish or exclude the diagnosis. Future prospective studies should determine whether using these models shortens diagnostic delay and improves care.

## Introduction

Pulmonary hypertension (PH) is a heterogeneous cardiopulmonary disorder defined by a mean pulmonary arterial pressure greater than 20 mm Hg at rest and may progress to right ventricular failure and death (1–3). Early recognition is important, yet diagnosis is often delayed because dyspnea, fatigue, and exercise intolerance are nonspecific (4,5).

Echocardiography is the primary noninvasive test for suspected PH, whereas right heart catheterization is required for definitive diagnosis and hemodynamic classification (1,6–8). The 12-lead electrocardiogram (ECG) is inexpensive and widely available, but established ECG findings have limited sensitivity for detecting PH (8,9).

Artificial intelligence (AI) can identify abnormalities in ECG waveforms that may not be apparent during routine interpretation. AI-enabled ECG models have detected ventricular dysfunction, atrial fibrillation, and other cardiovascular conditions (10–13). More recently, models have been developed to detect PH or elevated pulmonary artery pressure (14–18). However, their overall performance and potential clinical role remain uncertain.

We therefore conducted a systematic review and bivariate meta-analysis to quantify the performance of AI-enabled 12-lead ECG for detecting PH.

## Materials and methods

This systematic review and meta-analysis followed the Preferred Reporting Items for Systematic Reviews and Meta-Analyses (PRISMA) and PRISMA for Diagnostic Test Accuracy Studies guidelines (19,20). The protocol was prospectively registered in the International Prospective Register of Systematic Reviews (PROSPERO; CRD420251149532). This review used aggregate data from published studies and did not require institutional review board approval or informed consent. Completed reporting checklists are provided in Table A in S1 Appendix.

### Data sources and search strategy

ClinicalTrials.gov, the Cochrane Library, Google Scholar, Embase, MEDLINE, PubMed, Scopus, and Web of Science were searched from inception through May 2026. Controlled vocabulary and free-text terms related to artificial intelligence, machine learning, deep learning, electrocardiography, and pulmonary hypertension were combined. No language restrictions were applied. Database-specific search strategies are provided in Table B in S1 Appendix. Reference lists and citing articles of included studies and relevant reviews were also examined.

### Study selection

After deduplication, two reviewers (JE and AG) independently screened titles and abstracts and assessed potentially eligible full-text reports. Disagreements were resolved through discussion and, when necessary, adjudication by a third reviewer (HB). Reasons for full-text exclusion are provided in Table C in S1 Appendix. The study-selection process is summarized in Fig 1.

**Fig 1.**
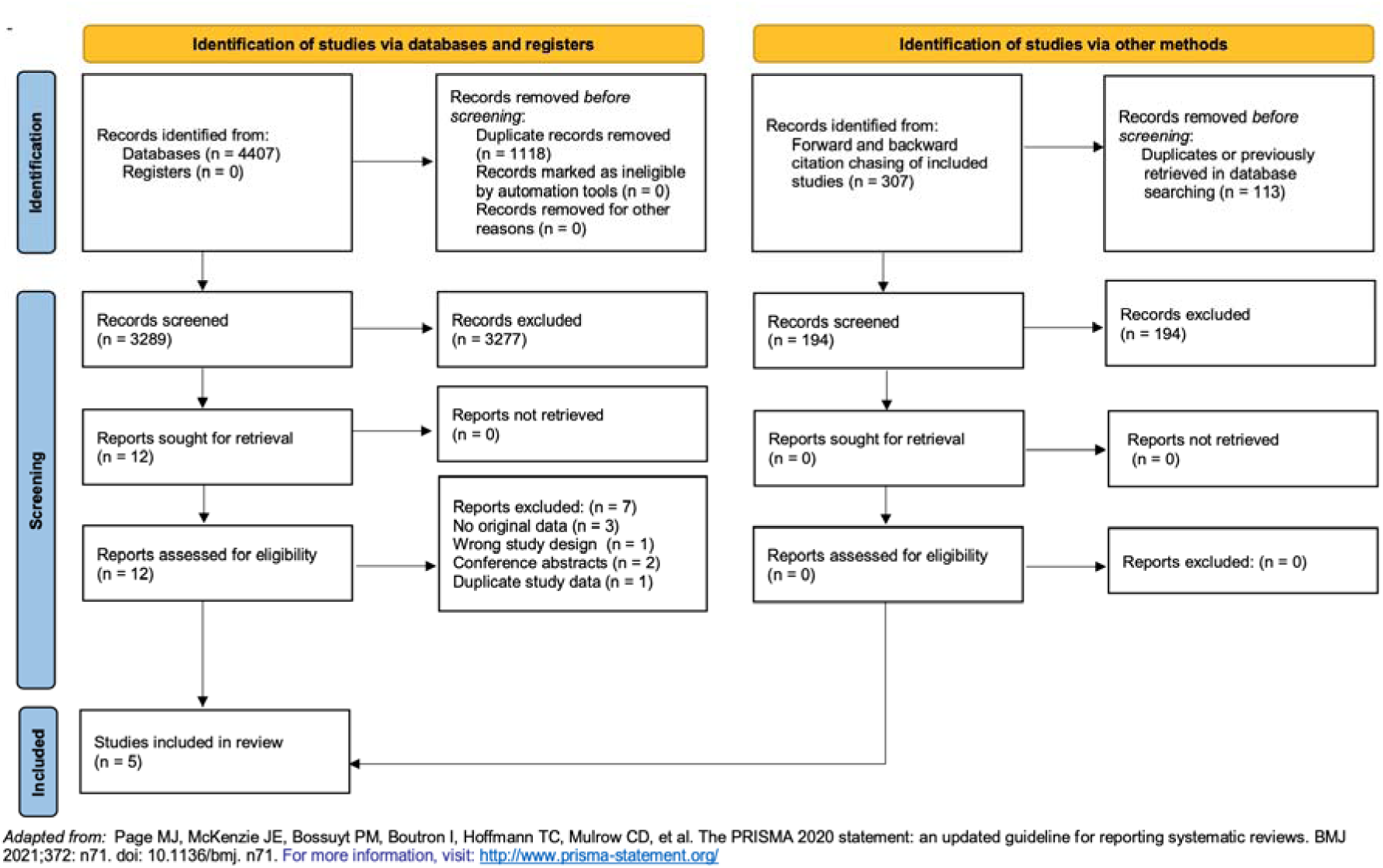
PRISMA flow diagram of study selection. The search identified 3,303 records; 14 duplicates were removed, 3,289 records were screened, 13 full-text reports were assessed, and five studies were included. PRISMA, Preferred Reporting Items for Systematic Reviews and Meta-Analyses.

### Eligibility criteria

Original peer-reviewed retrospective or prospective observational studies were eligible if they evaluated an AI or machine-learning algorithm applied to a standard 12-lead ECG for detecting PH or elevated pulmonary artery pressure. Eligible reference standards included right heart catheterization, echocardiographic assessment of pulmonary artery pressure or PH probability, or a prespecified hierarchical combination of these approaches.

Studies were required to report sensitivity and specificity or sufficient data to determine true-positive, false-positive, false-negative, and true-negative counts. Studies using wearable, single-lead, or other nonstandard ECG recordings were excluded, as were studies without an eligible reference standard, without extractable diagnostic performance data, or evaluating prognostic outcomes alone. Case reports, reviews, editorials, conference abstracts without sufficient data, and duplicate reports were also excluded.

### Data extraction

Two reviewers (JE and AG) independently extracted study-level data using a standardized form. Extracted variables included publication year, country, study design, clinical setting, sample size, study population, model architecture and inputs, ECG acquisition characteristics, PH definition, reference standard, interval between ECG and reference testing, operating threshold, validation strategy, sensitivity, specificity, and area under the receiver operating characteristic curve. True-positive, false-positive, false-negative, and true-negative counts were extracted or calculated from reported data. Disagreements were resolved through discussion and, when necessary, adjudication by a third reviewer (HB).

### Risk of bias assessment

Two reviewers (JE and AG) independently assessed risk of bias and applicability using the Quality Assessment of Diagnostic Accuracy Studies 2 (QUADAS-2) tool (21). Risk of bias was evaluated across patient selection, index test, reference standard, and flow and timing. Applicability concerns were evaluated for patient selection, index test, and reference standard. Each domain was classified as low, high, or unclear risk or concern. Disagreements were resolved through discussion and, when necessary, adjudication by a third reviewer (HB).

### Statistical analysis

Sensitivity and specificity were pooled using a bivariate random-effects model that accounted for within-study variability, between-study heterogeneity, and the correlation between sensitivity and specificity (22–24). Summary estimates are reported with 95% CIs. Positive and negative likelihood ratios and the diagnostic odds ratio were derived from the pooled model.

Study-specific and pooled sensitivity and specificity were displayed in forest plots. A summary receiver operating characteristic curve was generated with a summary operating point and 95% confidence and prediction regions. The area under the curve was estimated with its 95% CI. Heterogeneity was described using Cochran Q and I² statistics and interpreted with clinical and methodological differences across studies (25). Clinical utility was illustrated with a Fagan nomogram at an assumed pretest probability of 20% (26). Posttest probabilities following positive and negative AI-ECG results were calculated using the pooled likelihood ratios.

Leave-one-out analyses sequentially excluded each study and refitted the bivariate model. Publication bias was not formally assessed because fewer than 10 independent studies were available (27). Analyses were performed using Stata, version 18.0 (StataCorp LLC). Statistical tests were two-sided. A P value less than .10 was considered suggestive of heterogeneity; otherwise, P < .05 was considered statistically significant.

### Certainty of evidence

Certainty of evidence for pooled sensitivity and specificity was evaluated using the Grading of Recommendations Assessment, Development and Evaluation framework for diagnostic tests (28). Evidence was assessed for risk of bias, indirectness, inconsistency, imprecision, and publication bias and categorized as high, moderate, low, or very low certainty

## Results

### Study selection

The search identified 3,303 records, of which 14 duplicates were removed. Among 3,289 records screened by title and abstract, 3,276 were excluded and 13 full-text reports were assessed for eligibility. Eight full-text reports were excluded, leaving five studies for qualitative and quantitative synthesis (Fig 1). Reasons for full-text exclusion are provided in Table C in S1 Appendix.

### Study characteristics

The five included retrospective studies comprised 103,884 individuals and were published from 2020 through 2025 (14–18). Studies were conducted in South Korea, Taiwan, and the United States. Three studies contributed geographic external-validation cohorts, one contributed a held-out test cohort, and one used cross-validation.

The studies evaluated convolutional neural networks, deep neural networks, residual neural networks, and ensemble learning approaches. Four studies primarily analyzed raw 12-lead ECG waveforms, whereas Kwon et al incorporated demographic and conventional ECG measurements in addition to the ECG waveform (14). The interval between ECG acquisition and reference-standard testing ranged from 7 to 90 days.

Three studies used echocardiographic definitions of PH, whereas two used hierarchical reference standards incorporating right heart catheterization when available and echocardiography otherwise. Study characteristics are presented in Table 1. Reference standards, operating thresholds, and validation strategies are summarized in Table D in S1 Appendix, and diagnostic data are presented in Table E in S1 Appendix.

**Table 1.** Characteristics of studies included in the meta-analysis.

| Study | Country | N | Study population | Model inputs | AI model | ECG-reference interval |
| --- | --- | --- | --- | --- | --- | --- |
| Kwon et al, 2020 (14) | South Korea | 10,865 | Adults undergoing ECG and echocardiography at a community general hospital; geographic external-validation cohort | Raw 12-lead ECG plus age, sex, height, weight, and conventional ECG measurements | Ensemble CNN and DNN | ≤4 wk |
| Liu et al, 2022 (15) | Taiwan | 41,097 | Patients at Taipei Veterans General Hospital with an ECG-TTE pair obtained within 2 weeks | Raw 12-lead ECG waveform | Deep neural network | ≤2 wk |
| Aras et al, 2023 (16) | United States | 4,885 | Adults at UCSF undergoing RHC or echocardiography within 90 days of ECG; intermediate echocardiographic probability excluded | Raw 12-lead ECG waveform | 1-dimensional residual neural network | ≤90 d |
| DuBrock et al, 2024 (17) | United States | 30,301 | Adults at Vanderbilt University Medical Center with standard ECG and digital RHC or echocardiographic data; geographic external-validation cohort | Raw 12-lead ECG waveform | 1-dimensional CNN PH-EDA | ≤1 mo |
| Liu et al (Pang-Yen), 2025 (18) | Taiwan | 16,736 | Community-hospital patients with ECG within 7 days of echocardiography; geographic external-validation cohort | Raw 12-lead ECG waveform | 82-layer CNN | ≤7 d |

Abbreviations: AI, artificial intelligence; CNN, convolutional neural network; DNN, deep neural network; ECG, electrocardiogram; PH, pulmonary hypertension; RHC, right heart catheterization; TTE, transthoracic echocardiography.

### Diagnostic performance and clinical utility

Pooled sensitivity was 0.81 (95% CI, 0.79-0.82), and pooled specificity was 0.80 (95% CI, 0.76-0.83) (Fig 2). The area under the summary receiver operating characteristic curve was 0.85 (95% CI, 0.81-0.88) (Fig 3).

**Fig 2.**
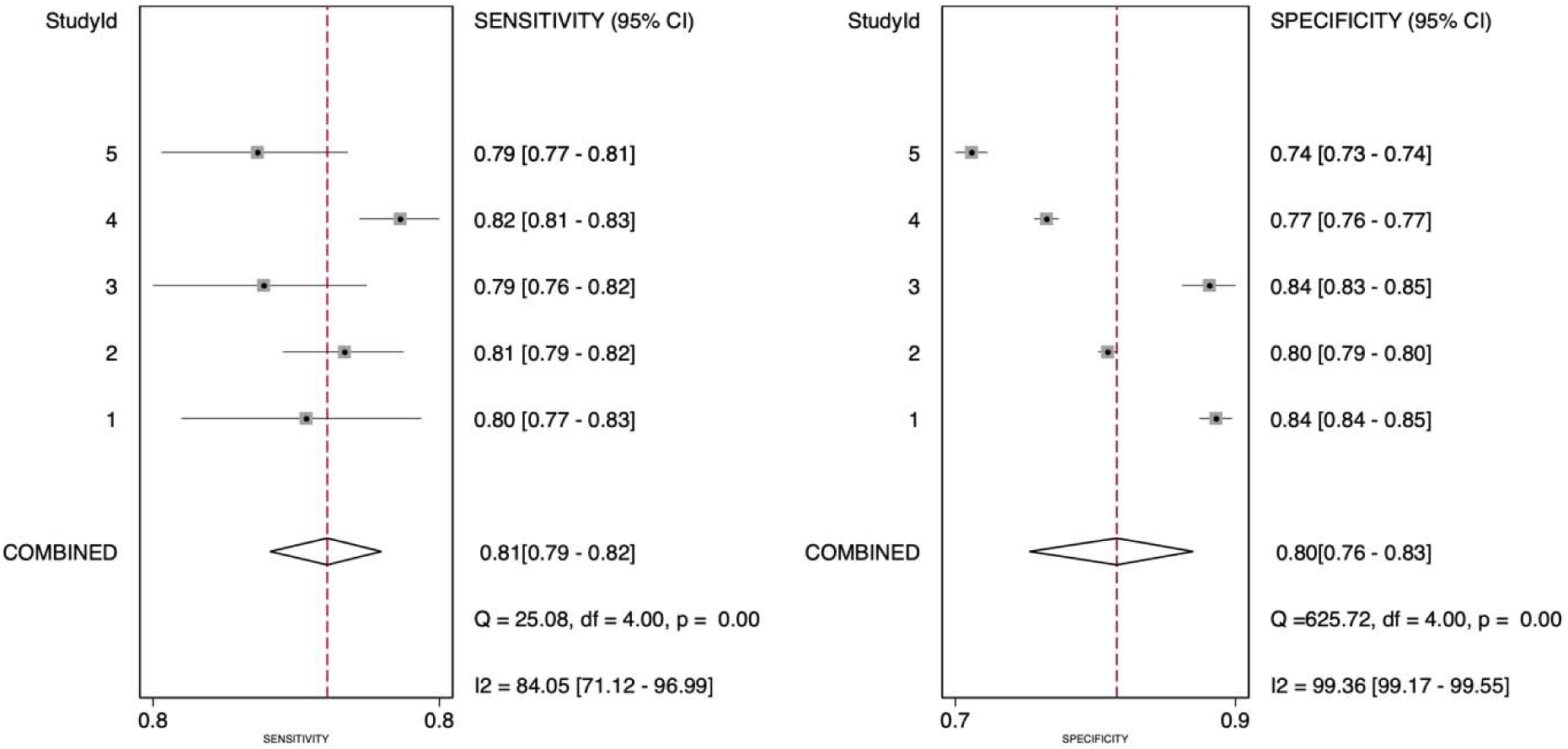
Sensitivity and specificity of AI-enabled electrocardiography for pulmonary hypertension. Forest plots show study-specific and pooled sensitivity and specificity. Squares represent study estimates, horizontal lines represent 95% CIs, diamonds represent pooled estimates, and dashed lines indicate pooled values. Study identifiers are 1, Kwon et al (14); 2, Liu et al (15); 3, Aras et al (16); 4, DuBrock et al (17); and 5, Liu et al (18).

**Fig 3.**
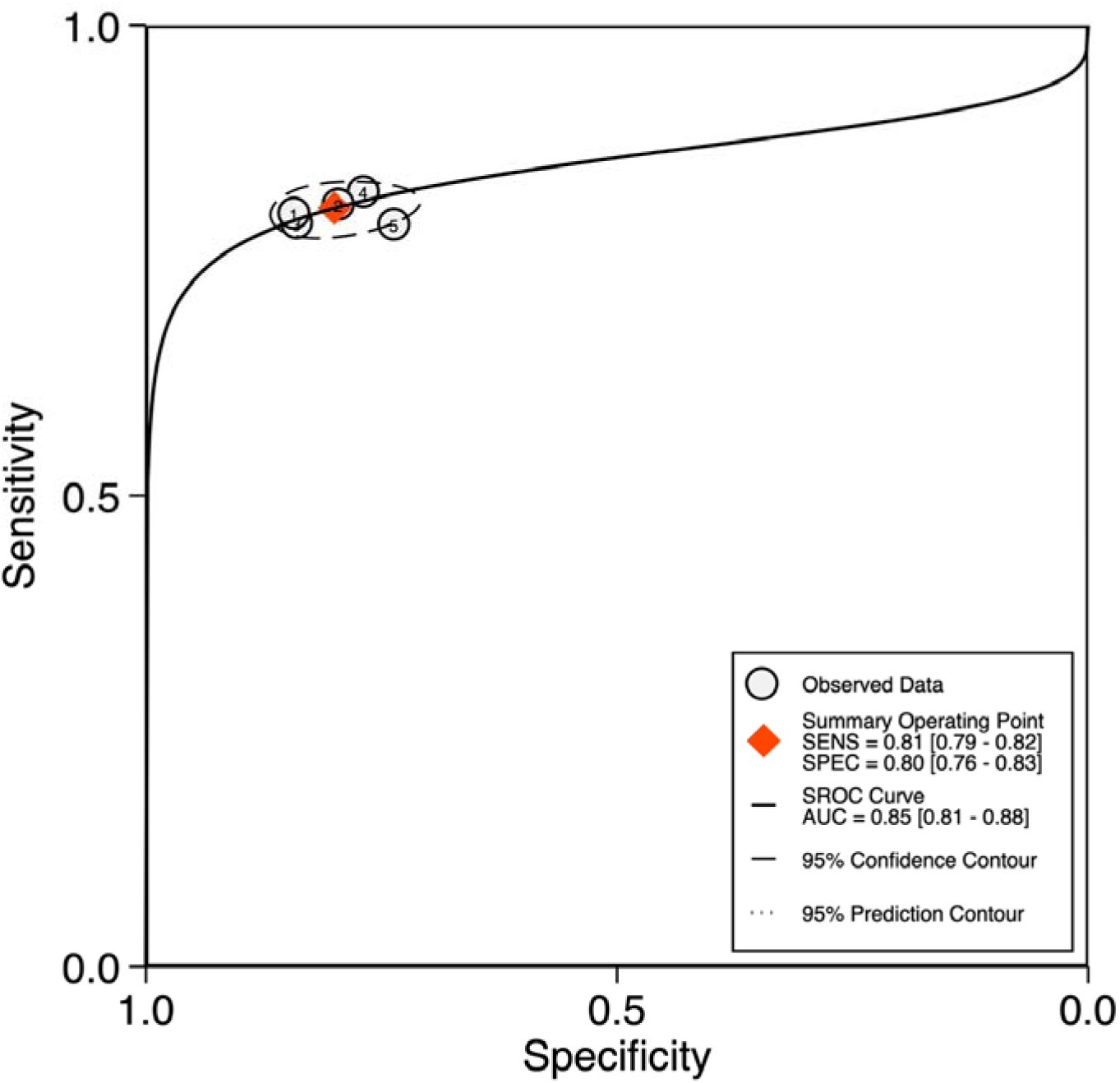
Summary receiver operating characteristic curve. Open circles represent individual studies; the orange diamond represents the summary operating point. The solid curve is the summary receiver operating characteristic curve, and contours indicate 95% confidence and prediction regions.

The pooled positive likelihood ratio was 4.03 (95% CI, 3.38-4.80), the negative likelihood ratio was 0.24 (95% CI, 0.23-0.26), and the diagnostic odds ratio was 16.58 (95% CI, 13.32-20.64). At an assumed pretest probability of 20%, a positive AI-ECG result increased the posttest probability of PH to approximately 50%, whereas a negative result reduced it to approximately 6% (Fig 4).

**Fig 4.**
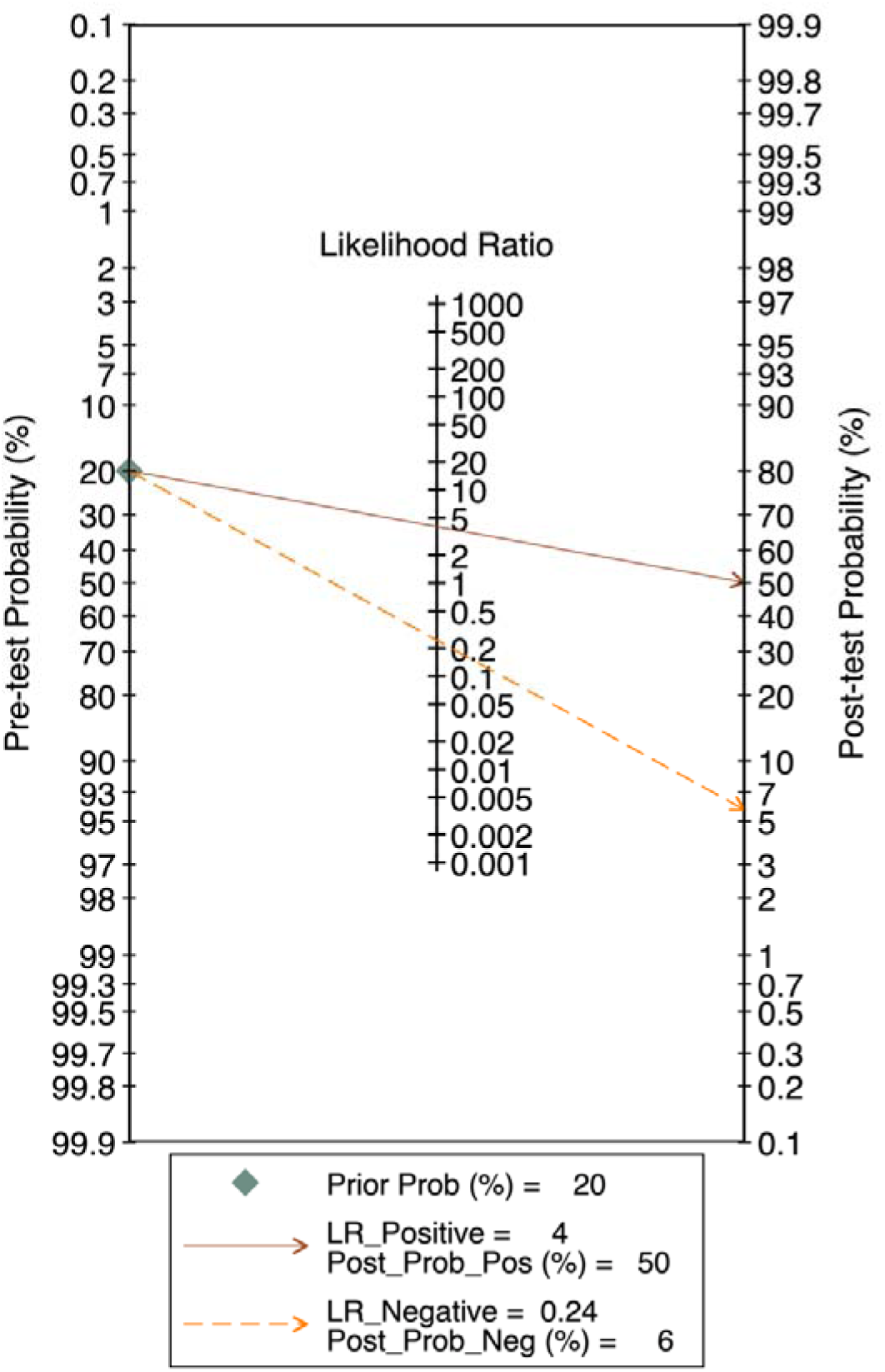
Fagan nomogram for AI-enabled electrocardiography. At a pretest probability of 20%, a positive AI-ECG result (positive likelihood ratio, 4.03) increased the posttest probability to approximately 50%, whereas a negative result (negative likelihood ratio, 0.24) reduced it to approximately 6%.

### Heterogeneity and sensitivity analyses

Heterogeneity was substantial. For sensitivity, I² was 84.1% (95% CI, 71.1%-97.0%; Q = 25.08; P < .001); for specificity, I² was 99.4% (95% CI, 99.2%-99.6%; Q = 625.72; P < .001) (Fig 2). Leave-one-out analyses did not materially alter the pooled estimates. Pooled sensitivity ranged from 0.80 to 0.81, pooled specificity from 0.79 to 0.81, the positive likelihood ratio from 3.80 to 4.33, and the negative likelihood ratio from 0.24 to 0.25 (Table G in S1 Appendix).

### Risk of bias and certainty of evidence

Risk-of-bias concerns were concentrated in patient selection, reference-standard classification, and flow and timing; index-test applicability was generally of low concern (Table 2). Certainty of evidence was rated moderate for sensitivity and low for specificity, with downgrading primarily for risk of bias and inconsistency (Table F in S1 Appendix).

**Table 2.** QUADAS-2 assessment of included studies.

| Study | Patient selection | Index test | Reference standard | Flow and timing | Patient selection applicability | Index test applicability | Reference standard applicability |
| --- | --- | --- | --- | --- | --- | --- | --- |
| Kwon et al, 2020 (14) | Low | Low | Unclear | Low | Low | Low | Low |
| Liu et al, 2022 (15) | Unclear | Low | Low | Unclear | Low | Low | Low |
| Aras et al, 2023 (16) | High | Low | Low | High | Unclear | Low | Low |
| DuBrock et al, 2024 (17) | Low | Low | Unclear | Low | Low | Low | Low |
| Liu et al, 2025 (18) | Low | Low | Low | Low | Low | Low | Low |

Risk of bias was assessed for patient selection, index test, reference standard, and flow and timing. Applicability concerns were assessed for patient selection, index test, and reference standard. QUADAS-2, Quality Assessment of Diagnostic Accuracy Studies 2.

## Discussion

In this systematic review and meta-analysis of five studies comprising 103,884 individuals, AI algorithms applied to standard 12-lead ECGs demonstrated good performance for detecting PH, with a pooled sensitivity of 0.81, specificity of 0.80, and summary area under the curve of 0.85. A negative AI-ECG substantially lowered, but did not eliminate, the probability of PH. Leave-one-out analyses yielded similar estimates, indicating that the findings were not driven by any single study. These results support AI-ECG as a potential case-finding or risk-enrichment tool rather than a stand-alone diagnostic test.

PH produces right ventricular pressure overload and remodeling that alter cardiac conduction and repolarization (8,9). Established ECG findings, including right-axis deviation and right ventricular hypertrophy, are often specific but insensitive (8,9). AI models interrogate the full waveform and may detect combinations of electrical features that are not evident during routine interpretation. Similar approaches have identified ventricular dysfunction and occult atrial fibrillation (10–13). The included studies extended this approach to PH across different populations and model architectures (14–18).

Clinical value will depend on where AI-ECG is placed in the diagnostic pathway. At a pretest probability of 20%, a positive result increased the posttest probability of PH to approximately 50%, whereas a negative result reduced it to approximately 6% (Fig 4). A positive result could identify patients who warrant echocardiography and, when indicated, right heart catheterization (1,6–8). A negative result could support triage but should not exclude PH when clinical suspicion remains high. The most plausible initial use is targeted case finding among patients with unexplained dyspnea, cardiopulmonary disease, or established PH risk factors.

Population-level screening would be less efficient. With a pooled specificity of 0.80, testing in low-prevalence populations could generate substantial false-positive downstream testing. Because posttest probability depends on baseline risk, targeted deployment in higher-risk populations is likely to provide greater clinical yield.

Reference standards and PH definitions varied across studies and likely contributed to the observed heterogeneity, particularly for specificity. Nevertheless, pooled estimates remained stable after sequential exclusion of each study (Table G in S1 Appendix). This supports the robustness of the summary estimates while indicating that real-world performance may vary with case mix, operating threshold, and diagnostic pathway.

Prospective multicenter studies should use prespecified PH definitions and thresholds, independent external validation, and transparent reporting (29–31). They should assess calibration, performance across PH subtypes and demographic groups, and robustness across ECG systems and clinical settings. Most importantly, future studies should determine whether AI-ECG shortens time to echocardiography or right heart catheterization, improves referral efficiency, or changes treatment and clinical outcomes.

This study has several limitations. First, PH definitions and reference standards varied across studies, contributing to clinical and methodological heterogeneity. Second, algorithms differed in architecture, ECG inputs, operating thresholds, and validation strategy. Third, aggregate study-level data precluded analyses by PH subtype, disease severity, demographic characteristics, comorbidities, or clinical setting. Fourth, diagnostic performance alone does not establish clinical utility, and the effects of AI-ECG on downstream testing, costs, diagnostic delay, treatment, and patient outcomes remain unknown.

AI-enabled electrocardiography demonstrated good performance for detecting PH and may support targeted case finding or risk enrichment. Current evidence is insufficient for routine clinical implementation. Prospective multicenter studies using standardized PH definitions and clinically relevant outcomes are needed to determine whether AI-ECG can improve diagnostic pathways and patient care.

## Data availability

All data used in this meta-analysis are reported in the article and S1 Appendix.

## Acknowledgments

## Supporting information

S1 Appendix. Supporting methods and analyses. Includes reporting guideline checklists, search strategies, excluded full-text reports, reference standards and validation strategies, diagnostic data, the GRADE evidence profile, and the leave-one-out sensitivity analysis.

