## Supplemental for "Clinical Usefulness of Artificial Intelligence–Enabled Electrocardiography for Pulmonary Hypertension Detection: A Meta-analysis Based on Pre- and Post-Test Probability"

### Contents

Table A. Reporting guideline checklists

Table B. Search strategies

Table C. Full-text reports excluded with reasons

Table D. Reference standards, operating thresholds, and validation strategies

Table E. Diagnostic data included in the meta-analysis

Table F. GRADE evidence profile

Table G. Leave-one-out sensitivity analysis

### Table A. Reporting Guideline Checklists

#### PRISMA 2020 Main Checklist

| **Topic** | **No.** | **Item** | **Location where item is reported** |
| --- | --- | --- | --- |
| **TITLE** |  |  |  |
| **Title** | 1 | Identify the report as a systematic review. | LN |
| **ABSTRACT** |  |  |  |
| **Abstract** | 2 | See the PRISMA 2020 for Abstracts checklist | Suppl.1 |
| **INTRODUCTION** |  |  |  |
| **Rationale** | 3 | Describe the rationale for the review in the context of existing knowledge. | LN |
| **Objectives** | 4 | Provide an explicit statement of the objective(s) or question(s) the review addresses. | LN |
| **METHODS** |  |  |  |
| **Eligibility criteria** | 5 | Specify the inclusion and exclusion criteria for the review and how studies were grouped for the syntheses. | LN |
| **Information sources** | 6 | Specify all databases, registers, websites, organisations, reference lists and other sources searched or consulted to identify studies. Specify the date when each source was last searched or consulted. | LN |
| **Search strategy** | 7 | Present the full search strategies for all databases, registers and websites, including any filters and limits used. | LN |
| **Selection process** | 8 | Specify the methods used to decide whether a study met the inclusion criteria of the review, including how many reviewers screened each record and each report retrieved, whether they worked independently, and if applicable, details of automation tools used in the process. | LN |
| **Data collection process** | 9 | Specify the methods used to collect data from reports, including how many reviewers collected data from each report, whether they worked independently, any processes for obtaining or confirming data from study investigators, and if applicable, details of automation tools used in the process. | LN |
| **Data items** | 10a | List and define all outcomes for which data were sought. Specify whether all results that were compatible with each outcome domain in each study were sought (e.g. for all measures, time points, analyses), and if not, the methods used to decide which results to collect. | LN |
|  | 10b | List and define all other variables for which data were sought (e.g. participant and intervention characteristics, funding sources). Describe any assumptions made about any missing or unclear information. | LN |
| **Study risk of bias assessment** | 11 | Specify the methods used to assess risk of bias in the included studies, including details of the tool(s) used, how many reviewers assessed each study and whether they worked independently, and if applicable, details of automation tools used in the process. | LN |
| **Effect measures** | 12 | Specify for each outcome the effect measure(s) (e.g. risk ratio, mean difference) used in the synthesis or presentation of results. | LN |
| **Synthesis methods** | 13a | Describe the processes used to decide which studies were eligible for each synthesis (e.g. tabulating the study intervention characteristics and comparing against the planned groups for each synthesis (item 5)). | LN |
|  | 13b | Describe any methods required to prepare the data for presentation or synthesis, such as handling of missing summary statistics, or data conversions. | LN |
|  | 13c | Describe any methods used to tabulate or visually display results of individual studies and syntheses. | LN |
|  | 13d | Describe any methods used to synthesize results and provide a rationale for the choice(s). If meta-analysis was performed, describe the model(s), method(s) to identify the presence and extent of statistical heterogeneity, and software package(s) used. | LN |
|  | 13e | Describe any methods used to explore possible causes of heterogeneity among study results (e.g. subgroup analysis, meta-regression). | LN |
|  | 13f | Describe any sensitivity analyses conducted to assess robustness of the synthesized results. | LN |
| **Reporting bias assessment** | 14 | Describe any methods used to assess risk of bias due to missing results in a synthesis (arising from reporting biases). | LN |
| **Certainty assessment** | 15 | Describe any methods used to assess certainty (or confidence) in the body of evidence for an outcome. | LN |
| **RESULTS** |  |  |  |
| **Study selection** | 16a | Describe the results of the search and selection process, from the number of records identified in the search to the number of studies included in the review, ideally using a flow diagram. | LN |
|  | 16b | Cite studies that might appear to meet the inclusion criteria, but which were excluded, and explain why they were excluded. | Suppl. Table 3 |
| **Study characteristics** | 17 | Cite each included study and present its characteristics. | LN |
| **Risk of bias in studies** | 18 | Present assessments of risk of bias for each included study. | LN |
| **Results of individual studies** | 19 | For all outcomes, present, for each study: (a) summary statistics for each group (where appropriate) and (b) an effect estimate and its precision (e.g. confidence/credible interval), ideally using structured tables or plots. | LN |
| **Results of syntheses** | 20a | For each synthesis, briefly summarise the characteristics and risk of bias among contributing studies. | LN |
|  | 20b | Present results of all statistical syntheses conducted. If meta-analysis was done, present for each the summary estimate and its precision (e.g. confidence/credible interval) and measures of statistical heterogeneity. If comparing groups, describe the direction of the effect. | LN |
|  | 20c | Present results of all investigations of possible causes of heterogeneity among study results. | LN |
|  | 20d | Present results of all sensitivity analyses conducted to assess the robustness of the synthesized results. | LN |
| **Reporting biases** | 21 | Present assessments of risk of bias due to missing results (arising from reporting biases) for each synthesis assessed. | LN |
| **Certainty of evidence** | 22 | Present assessments of certainty (or confidence) in the body of evidence for each outcome assessed. | LN |
| **DISCUSSION** |  |  |  |
| **Discussion** | 23a | Provide a general interpretation of the results in the context of other evidence. | LN |
|  | 23b | Discuss any limitations of the evidence included in the review. | LN |
|  | 23c | Discuss any limitations of the review processes used. | LN |
|  | 23d | Discuss implications of the results for practice, policy, and future research. | LN |
| **OTHER INFORMATION** |  |  |  |
| **Registration and protocol** | 24a | Provide registration information for the review, including register name and registration number, or state that the review was not registered. | LN |
|  | 24b | Indicate where the review protocol can be accessed, or state that a protocol was not prepared. | LN |
|  | 24c | Describe and explain any amendments to information provided at registration or in the protocol. | N/A |
| **Support** | 25 | Describe sources of financial or non-financial support for the review, and the role of the funders or sponsors in the review. | LN |
| **Competing interests** | 26 | Declare any competing interests of review authors. | LN |
| **Availability of data, code and other materials** | 27 | Report which of the following are publicly available and where they can be found: template data collection forms; data extracted from included studies; data used for all analyses; analytic code; any other materials used in the review. | LN |

From: Page MJ, McKenzie JE, Bossuyt PM, Boutron I, Hoffmann TC, Mulrow CD, et al. The PRISMA 2020 statement: an updated guideline for reporting systematic reviews. MetaArXiv. 2020, September 14. DOI: 10.31222/osf.io/v7gm2. For more information, visit: www.prisma-statement.org

#### PRISMA Abstract Checklist

| **Topic** | **No.** | **Item** | **Reported?** |
| --- | --- | --- | --- |
| **TITLE** |  |  |  |
| **Title** | 1 | Identify the report as a systematic review. | Yes |
| **BACKGROUND** |  |  |  |
| **Objectives** | 2 | Provide an explicit statement of the main objective(s) or question(s) the review addresses. | Yes |
| **METHODS** |  |  |  |
| **Eligibility criteria** | 3 | Specify the inclusion and exclusion criteria for the review. | Yes |
| **Information sources** | 4 | Specify the information sources (e.g. databases, registers) used to identify studies and the date when each was last searched. | Yes |
| **Risk of bias** | 5 | Specify the methods used to assess risk of bias in the included studies. | Yes |
| **Synthesis of results** | 6 | Specify the methods used to present and synthesize results. | Yes |
| **RESULTS** |  |  |  |
| **Included studies** | 7 | Give the total number of included studies and participants and summarise relevant characteristics of studies. | Yes |
| **Synthesis of results** | 8 | Present results for main outcomes, preferably indicating the number of included studies and participants for each. If meta-analysis was done, report the summary estimate and confidence/credible interval. If comparing groups, indicate the direction of the effect (i.e. which group is favoured). | Yes |
| **DISCUSSION** |  |  |  |
| **Limitations of evidence** | 9 | Provide a brief summary of the limitations of the evidence included in the review (e.g. study risk of bias, inconsistency and imprecision). | Yes |
| **Interpretation** | 10 | Provide a general interpretation of the results and important implications. | Yes |
| **OTHER** |  |  |  |
| **Funding** | 11 | Specify the primary source of funding for the review. | Yes |
| **Registration** | 12 | Provide the register name and registration number. | Yes |

| **Section/topic** | **#** | **PRISMA-DTA Checklist Item** | **Reported on page #** |
| --- | --- | --- | --- |
| **TITLE / ABSTRACT** | | |  |
| Title | 1 | Identify the report as a systematic review (+/- meta-analysis) of diagnostic test accuracy (DTA) studies. |  |
| Abstract | 2 | Abstract: See PRISMA-DTA for abstracts. |  |
| **INTRODUCTION** | | |  |
| Rationale | 3 | Describe the rationale for the review in the context of what is already known. |  |
| Clinical role of index test | D1 | State the scientific and clinical background, including the intended use and clinical role of the index test, and if applicable, the rationale for minimally acceptable test accuracy (or minimum difference in accuracy for comparative design). |  |
| Objectives | 4 | Provide an explicit statement of question(s) being addressed in terms of participants, index test(s), and target condition(s). |  |
| **METHODS** | | |  |
| Protocol and registration | 5 | Indicate if a review protocol exists, if and where it can be accessed (e.g., Web address), and, if available, provide registration information including registration number. |  |
| Eligibility criteria | 6 | Specify study characteristics (participants, setting, index test(s), reference standard(s), target condition(s), and study design) and report characteristics (e.g., years considered, language, publication status) used as criteria for eligibility, giving rationale. |  |
| Information sources | 7 | Describe all information sources (e.g., databases with dates of coverage, contact with study authors to identify additional studies) in the search and date last searched. |  |
| Search | 8 | Present full search strategies for all electronic databases and other sources searched, including any limits used, such that they could be repeated. |  |
| Study selection | 9 | State the process for selecting studies (i.e., screening, eligibility, included in systematic review, and, if applicable, included in the meta-analysis). |  |
| Data collection process | 10 | Describe method of data extraction from reports (e.g., piloted forms, independently, in duplicate) and any processes for obtaining and confirming data from investigators. |  |
| **Section/topic** | **#** | **PRISMA-DTA Checklist Item** | **Reported on page #** |
| Definitions for data extraction | 11 | Provide definitions used in data extraction and classifications of target condition(s), index test(s), reference standard(s) and other characteristics (e.g. study design, clinical setting). |  |
| Risk of bias and applicability | 12 | Describe methods used for assessing risk of bias in individual studies and concerns regarding the applicability to the review question. |  |
| Diagnostic accuracy measures | 13 | State the principal diagnostic accuracy measure(s) reported (e.g. sensitivity, specificity) and state the unit of assessment (e.g. per-patient, per-lesion). |  |
| Synthesis of results | 14 | Describe methods of handling data, combining results of studies and describing variability between studies. This could include, but is not limited to: a) handling of multiple definitions of target condition. b) handling of multiple thresholds of test positivity, c) handling multiple index test readers, d) handling of indeterminate test results, e) grouping and comparing tests, f) handling of different reference standards |  |
| Meta-analysis | D2 | Report the statistical methods used for meta-analyses, if performed. |  |
| Additional analyses | 16 | Describe methods of additional analyses (e.g., sensitivity or subgroup analyses, meta-regression), if done, indicating which were pre-specified. |  |
| **RESULTS** | | | |
| Study selection | 17 | Provide numbers of studies screened, assessed for eligibility, included in the review (and included in meta-analysis, if applicable) with reasons for exclusions at each stage, ideally with a flow diagram. |  |
| Study characteristics | 18 | For each included study provide citations and present key characteristics including: a) participant characteristics (presentation, prior testing), b) clinical setting, c) study design, d) target condition definition, e) index test, f) reference standard, g) sample size, h) funding sources |  |
| Risk of bias and applicability | 19 | Present evaluation of risk of bias and concerns regarding applicability for each study. |  |
| Results of individual studies | 20 | For each analysis in each study (e.g. unique combination of index test, reference standard, and positivity threshold) report 2x2 data (TP, FP, FN, TN) with estimates of diagnostic accuracy and confidence intervals, ideally with a forest or receiver operator characteristic (ROC) plot. |  |
| Synthesis of results | 21 | Describe test accuracy, including variability; if meta-analysis was done, include results and confidence intervals. |  |

Page 1 of 2

| **Section/topic** | **#** | **PRISMA-DTA Checklist Item** | **Reported on page #** |
| --- | --- | --- | --- |
| Additional analysis | 23 | Give results of additional analyses, if done (e.g., sensitivity or subgroup analyses, meta-regression; analysis of index test: failure rates, proportion of inconclusive results, adverse events). |  |
| **DISCUSSION** | | |  |
| Summary of evidence | 24 | Summarize the main findings including the strength of evidence. |  |
| Limitations | 25 | Discuss limitations from included studies (e.g. risk of bias and concerns regarding applicability) and from the review process (e.g. incomplete retrieval of identified research). |  |
| Conclusions | 26 | Provide a general interpretation of the results in the context of other evidence. Discuss implications for future research and clinical practice (e.g. the intended use and clinical role of the index test). |  |
| **FUNDING** | | |  |
| Funding | 27 | For the systematic review, describe the sources of funding and other support and the role of the funders. |  |

*Adapted From:* McInnes MDF, Moher D, Thombs BD, McGrath TA, Bossuyt PM, The PRISMA-DTA Group (2018). Preferred Reporting Items for a Systematic Review and Meta-analysis of Diagnostic Test Accuracy Studies: The PRISMA-DTA Statement. JAMA. 2018 Jan 23;319(4):388-396. doi: 10.1001/jama.2017.19163.

### Table B. Search Strategies

#### Ovid Embase

1	exp pulmonary hypertension/

2	((pulmonar* or lung or portopulmonar* or porto-pulmonar*) adj3 (hypertens* or hemangiomatos* or haemangiomatos* or venoocclusive or high-blood-pressure* or elevate*-blood-pressure*)).tw,kf.

3	(cor pulmonale or corpulmonale or right heart failure* or right heart disease* or eisenmenger*).tw,kf.

4	((pulmonar* or lung) adj3 vascular adj3 (disease* or disorder* or failure)).tw,kf.

5	or/1-4

6	exp electrocardiography/

7	(electrocardiograph* or electro-cardiograph* or electromyocardiograph* or polycardiograph* or Cardiograph* or Electrocardiogram* or Electrocardiograph* or ECG or EKG or ECGs or EKGs or Vectorcardiograph* or Vectercardiograph* or polarcardiograph* or Body-Surface-Mapping*).tw,kf.

8	6 or 7

9	5 and 8

10	exp artificial intelligence/

11	exp "feature learning (machine learning)"/

12	exp computer assisted diagnosis/

13	((artificial or computat* or computer* or machine or deep or transfer or hierarchical) adj3 (intelligence* or learning* or reasoning* or algorithm*)).tw,kf.

14	(neural network* or random forest* or decision tree* or knowledge representation* or computer vision system* or computer reasoning* or perceptron* or connectionist model* or expert system* or computer assisted).tw,kf.

15	((ECG or EKG or electrocardiograph* or electrocardiogram* or diagnos*) adj3 algorithm*).tw,kf.

16	AI.ti,ab.

17	or/10-16

18	9 and 17

#### Ovid MEDLINE(R) ALL

1	exp Hypertension, Pulmonary/

2	((pulmonar* or lung or portopulmonar* or porto-pulmonar*) adj3 (hypertens* or hemangiomatos* or haemangiomatos* or venoocclusive or high-blood-pressure* or elevate*-blood-pressure*)).tw,kf.	67508

3	(cor-pulmonale or corpulmonale or eisenmenger*).tw,kf.

4	(Right-heart adj3 (disease* or failure* or disorder*)).tw,kf.

5	((pulmonar* or lung) adj3 vascular adj3 (disease* or disorder* or failure)).tw,kf

6	or/1-5

7	exp Electrocardiography/

8	(electrocardiograph* or electro-cardiograph* or electromyocardiograph* or polycardiograph* or Cardiograph* or Electrocardiogram* or Electrocardiograph* or ECG or EKG or ECGs or EKGs or Vectorcardiograph* or Vectercardiograph* or polarcardiograph* or Body-Surface-Mapping*).tw,kf.

9	7 or 8

10	6 and 9

11	exp Artificial Intelligence/

12	exp Diagnosis, Computer-Assisted/

13	((artificial or computat* or computer* or machine or deep or transfer or hierarchical) adj3 (intelligence* or learning* or reasoning* or algorithm*)).tw,kf.

14	(neural network* or random forest* or decision tree* or knowledge representation* or computer vision system* or computer reasoning* or perceptron* or connectionist model* or expert system* or computer assisted).tw,kf.

15	((ECG or EKG or electrocardiograph* or electrocardiogram* or diagnos*) adj3 algorithm*).tw,kf.	16710

16	AI.ti,ab.

17	or/11-16

18	10 and 17

#### Web of Science Core Collection

#1 TS=((artificial or computat* or computer* or machine or deep or transfer or hierarchical) near/3 (intelligence* or learning* or reasoning* or algorithm*)) or TS=("neural network*" or "random forest*" or "decision tree*" or "knowledge representation*" or "computer vision system*" or "computer reasoning*" or perceptron* or "connectionist model*" or "expert system*" or "computer assisted" or AI) or TS=((ECG or EKG or electrocardiograph* or electrocardiogram* or diagnos*) near/3 algorithm*)

#2 TS=(electrocardiograph* or electro-cardiograph* or electromyocardiograph* or polycardiograph* or Cardiograph* or Electrocardiogram* or Electrocardiograph* or ECG or EKG or ECGs or EKGs or Vectorcardiograph* or Vectercardiograph* or polarcardiograph* or Body-Surface-Mapping*)

#3 TS=((pulmonar* or lung or portopulmonar* or porto-pulmonar*) near/3 (hypertens* or hemangiomatos* or haemangiomatos* or venoocclusive or high-blood-pressure* or elevate*-blood-pressure*)) or TS=(cor-pulmonale or corpulmonale or eisenmenger*) or TS=(Right-heart near/3 (disease* or failure* or disorder*)) or TS=((pulmonar* or lung) near/3 vascular near/3 (disease* or disorder* or failure))

#4 #1 and #2 and #3

The Core Collection included in this review includes:

1.	Science Citation Index Expanded (1900 - Data Searched)

2.	Social Sciences Citation Index (1900 - Date Searched)

3.	Art & Humanities - (1975 - Date Searched)

4.	Conference Proceedings Citation Index - Science (1991 - Date Searched)

5.	Conference Proceedings Citation Index - Social Sciences and Humanities (1991 - Date Searched)

6.	Book Citation Index - Science (2005 - Date Searched)

7.	Book Citation Index - Social Sciences and Humanities ( 2005 - Date Searched)

8.	Emerging Source Citation Index - (2018 - Date Searched)

9.	Current Chemical Reactions (1985 - Date Searched)

10.	Index Chemicus (1993 - Date Searched)

#### Cochrane Library

#1 (electrocardiograph* or electro-cardiograph* or electromyocardiograph* or polycardiograph* or Cardiograph* or Electrocardiogram* or Electrocardiograph* or ECG or EKG or ECGs or EKGs or Vectorcardiograph* or Vectercardiograph* or polarcardiograph* or Body-Surface-Mapping*):ti,ab

#2 ((pulmonar* or lung or portopulmonar* or porto-pulmonar*) near/3 (hypertens* or hemangiomatos* or haemangiomatos* or venoocclusive or high blood pressure* or elevate* blood pressure*)):ti,ab or (cor-pulmonale or corpulmonale or eisenmenger*):ti,ab or (Right-heart near/3 (disease* or failure* or disorder*)):ti,ab or ((pulmonar* or lung) near/3 vascular near/3 (disease* or disorder* or failure)):ti,ab

#3 ((artificial or computat* or computer* or machine or deep or transfer or hierarchical) near/3 (intelligence* or learning* or reasoning* or algorithm*)):ti,ab or (neural network* or random forest* or decision tree* or knowledge representation* or computer vision system* or computer reasoning* or perceptron* or connectionist model* or expert system* or computer assisted or AI):ti,ab or ((ECG or EKG or electrocardiograph* or electrocardiogram* or diagnos*) near/3 algorithm*):ti,ab

#4 #1 and #2 and #3

The Cochrane Library database includes:

1.	Cochrane Database of Systematic Reviews

2.	Cochrane Central Register of Controlled Trials

3.	Cochrane Clinical Answers

#### Scopus

( TITLE-ABS-KEY ( ( pulmonar* OR lung OR portopulmonar* OR porto-pulmonar* ) W/3 ( hypertens* OR hemangiomatos* OR haemangiomatos* OR venoocclusive OR high-blood-pressure* OR elevate*-blood-pressure* ) ) OR TITLE-ABS-KEY ( cor-pulmonale OR corpulmonale OR eisenmenger* ) OR TITLE-ABS-KEY ( Right-heart W/3 ( disease* OR failure* OR disorder* ) ) OR TITLE-ABS-KEY ( ( pulmonar* OR lung ) W/3 vascular W/3 ( disease* OR disorder* OR failure ) ) ) AND ( TITLE-ABS-KEY ( electrocardiograph* OR electro-cardiograph* OR electromyocardiograph* OR polycardiograph* OR Cardiograph* OR Electrocardiogram* OR Electrocardiograph* OR ECG OR EKG OR ECGs OR EKGs OR Vectorcardiograph* OR Vectercardiograph* OR polarcardiograph* OR Body-Surface-Mapping* ) ) AND ( TITLE-ABS-KEY ( ( artificial OR computat* OR computer* OR machine OR deep OR transfer OR hierarchical ) W/3 ( intelligence* OR learning* OR reasoning* OR algorithm* ) ) OR TITLE-ABS-KEY ( "neural network*" OR "random forest*" OR "decision tree*" OR "knowledge representation*" OR "computer vision system*" OR "computer reasoning*" OR perceptron* OR "connectionist model*" OR "expert system*" OR "computer assisted" OR AI ) OR TITLE-ABS-KEY ( ( ECG OR EKG OR electrocardiograph* OR electrocardiogram* OR diagnos* ) W/3 algorithm* ) )

#### Google Scholar (via Harzing’s Publish or Perish Edition: 8.19.5300.9453)

pulmonary hypertension electrocardiogram artificial intelligence

### Table C. Full-Text Reports Excluded With Reasons

| **First author** | **Year** | **Title** | **Source** | **Reason for exclusion** |
| --- | --- | --- | --- | --- |
| ClinicalTrials.gov registry | 2025 | A Deep-Learning-Enabled Electrocardiogram for Detecting Pulmonary Hypertension: A Randomized Controlled Trial | ClinicalTrials.gov | No original results |
| DuBrock | 2022 | Early Detection of Pulmonary Arterial Hypertension Using a Pulmonary Hypertension Electrocardiogram Algorithm | Am J Respir Crit Care Med | Duplicate study data |
| Huang | 2025 | AI-Enhanced ECG for Pulmonary Hypertension Detection: Integrating Machine Learning and Deep Learning Approaches | Eur Heart J | Conference abstract without sufficient data |
| Kaihara | 2025 | Can an AI-Powered Smartphone App Estimate Moderate Pulmonary Hypertension by Taking a 12-Lead ECG of Japanese Patients? | Eur Heart J | Conference abstract without sufficient data |
| Kishikawa | 2025 | An Ensemble Learning Model for Detection of Pulmonary Hypertension Using Electrocardiogram, Chest X-Ray, and Brain Natriuretic Peptide | Eur Heart J Digit Health | Wrong study design |
| Langlais | 2023 | Overcoming Diagnostic Delays in Pulmonary Hypertension With Deep-Learning ECG Analysis | J Card Fail | No original data |
| Van der Bijl | 2022 | Using Deep Learning to Diagnose Pulmonary Hypertension | Eur Heart J Cardiovasc Imaging | No original data |
| Nazareth | 2026 | Diagnostic Performance of Artificial Intelligence-Enabled Electrocardiography for Pulmonary Hypertension: A Systematic Review and Meta-analysis | Eur Heart J Digit Health | Systematic review; no original data |

### Table D. Reference Standards, Operating Thresholds, and Validation Strategies

| **Study** | **PH reference standard** | **Operating threshold** | **Validation strategy** |
| --- | --- | --- | --- |
| Kwon et al, 2020 | Echocardiographic pulmonary artery systolic pressure >50 mm Hg | Derivation cutoff selected to achieve 80% sensitivity | Internal validation at Hospital A and geographic external validation at Hospital B; Hospital B used in the primary meta-analysis |
| Liu et al, 2022 | Echocardiographic pulmonary artery systolic pressure >50 mm Hg | Cross-validation threshold | Ten-fold cross-validation in the primary Taiwanese cohort; a small Japanese external cohort was also reported |
| Aras et al, 2023 | RHC mPAP >20 mm Hg when available; otherwise echocardiographic TR velocity >3.4 m/s | Threshold optimized using the F1 score in the development data set | Held-out test cohort from the same health system; no geographic external validation |
| DuBrock et al, 2024 | Hierarchical RHC/echocardiographic definition using mPAP >20 mm Hg or TR velocity >3.4 m/s | Fixed probability threshold of 0.308 | Testing at Mayo Clinic and geographic external validation at Vanderbilt; Vanderbilt used in the primary meta-analysis |
| Liu et al (Pang-Yen), 2025 | Echocardiographic pulmonary artery systolic pressure >50 mm Hg | Maximum Youden index threshold | Internal validation at Hospital A and geographic external validation at Hospital B; Hospital B ECG-only cohort used in the primary meta-analysis |

Abbreviations: ECG, electrocardiogram; mPAP, mean pulmonary arterial pressure; PH, pulmonary hypertension; RHC, right heart catheterization; TR, tricuspid regurgitation.

### Table E. Diagnostic Data Included in the Meta-analysis

| **Study** | **Cohort** | **N** | **PH+** | **PH-** | **TP** | **FN** | **TN** | **FP** | **Sensitivity** | **Specificity** |
| --- | --- | --- | --- | --- | --- | --- | --- | --- | --- | --- |
| Kwon et al, 2020 | Hospital B external | 10,865 | 757 | 10108 | 606 | 151 | 8521 | 1587 | 0.801 | 0.843 |
| Liu et al, 2022 | Taipei cross-validation | 41,097 | 2824 | 38273 | 2287 | 537 | 30465 | 7808 | 0.810 | 0.796 |
| Aras et al, 2023 | UCSF held-out test | 4,885 | 982 | 3903 | 776 | 206 | 3279 | 624 | 0.790 | 0.840 |
| DuBrock et al, 2024 | Vanderbilt external | 30,301 | 6045 | 24256 | 4977 | 1068 | 18665 | 5591 | 0.823 | 0.770 |
| Liu et al (Pang-Yen), 2025 | Hospital B external ECG-only | 16,736 | 1292 | 15444 | 1019 | 273 | 11382 | 4062 | 0.789 | 0.737 |

Counts were reconstructed from reported disease-positive and disease-negative denominators and reported sensitivity and specificity estimates. Abbreviations: FN, false negative; FP, false positive; PH, pulmonary hypertension; TN, true negative; TP, true positive.

### Table F. GRADE Evidence Profile

| **Outcome** | **Studies** | **Study design** | **Risk of bias** | **Indirectness** | **Inconsistency** | **Imprecision** | **Publication bias** | **Certainty** |
| --- | --- | --- | --- | --- | --- | --- | --- | --- |
| Sensitivity | 5 | Observational diagnostic accuracy studies | Serious | Not serious | Serious | Not serious | Not assessed | Moderate |
| Specificity | 5 | Observational diagnostic accuracy studies | Very serious | Not serious | Very serious | Not serious | Not assessed | Low |

Publication bias was not formally assessed because fewer than 10 independent studies were available. GRADE indicates Grading of Recommendations Assessment, Development and Evaluation.

### Table G. Leave-One-Out Sensitivity Analysis

| **Excluded study** | **Pooled sensitivity (95% CI)** | **Pooled specificity (95% CI)** | **LR+** | **LR-** | **DOR** |
| --- | --- | --- | --- | --- | --- |
| Kwon et al, 2020 | 0.806 (0.791-0.821) | 0.788 (0.748-0.823) | 3.801 | 0.246 | 15.469 |
| Liu et al, 2022 | 0.804 (0.786-0.820) | 0.801 (0.753-0.842) | 4.038 | 0.245 | 16.464 |
| Aras et al, 2023 | 0.809 (0.795-0.823) | 0.789 (0.747-0.825) | 3.835 | 0.242 | 15.865 |
| DuBrock et al, 2024 | 0.801 (0.788-0.813) | 0.807 (0.763-0.844) | 4.151 | 0.247 | 16.823 |
| Liu et al, 2025 | 0.806 (0.788-0.822) | 0.814 (0.782-0.842) | 4.327 | 0.239 | 18.130 |

The overall model yielded sensitivity of 0.806 (95% CI, 0.792-0.819), specificity of 0.800 (95% CI, 0.762-0.833), LR+ of 4.028, LR- of 0.243, and DOR of 16.577. Abbreviations: CI, confidence interval; DOR, diagnostic odds ratio; LR, likelihood ratio.
